# Self-supervised Learning for Chest CT - Training Strategies and Effect on Downstream Applications

**DOI:** 10.1101/2024.02.01.24302144

**Authors:** Amara Tariq, Bhavik N. Patel, Imon Banerjee

## Abstract

Self-supervised pretraining can reduce the amount of labeled training data needed by pre-learning fundamental visual characteristics of the medical imaging data. In this study, we investigate several self-supervised training strategies for chest computed tomography exams and their effects of downstream applications. we bench-mark five well-known self-supervision strategies (masked image region prediction, next slice prediction, rotation prediction, flip prediction and denoising) on 15M chest CT slices collected from four sites of Mayo Clinic enterprise. These models were evaluated for two downstream tasks on public datasets; pulmonary embolism (PE) detection (classification) and lung nodule segmentation. Image embeddings generated by these models were also evaluated for prediction of patient age, race, and gender to study inherent biases in models’ understanding of chest CT exams. Use of pretraining weights, especially masked regions prediction based weights, improved performance and reduced computational effort needed for downstream tasks compared to task-specific state-of-the-art (SOTA) models. Performance improvement for PE detection was observed for training dataset sizes as large as 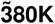 with maximum gain of 5% over SOTA. Segmentation model initialized with pretraining weights learned twice as fast as randomly initialized model. While gender and age predictors built using self-supervised training weights showed no performance improvement over randomly initialized predictors, the race predictor experienced a 10% performance boost when using self-supervised training weights. We released models and weights under open-source academic license. These models can then be finetuned with limited task-specific annotated data for a variety of downstream imaging tasks thus accelerating research in biomedical imaging informatics.

## 1. Introduction

**D**EEP learning models have been proposed and trained for high-dimensional medical imaging data, such as CT or MRI volumes, to target a variety of image interpretation tasks, and have demonstrated diagnostic accuracy comparable to that of human experts [11], [18], [21], [24]. While it provides promising solutions, the current success has been largely dominated by supervised learning frameworks, which typically require large-scale labeled datasets to achieve high performance. If the size of training data is limited, the deep learning models often suffer from over-fitting (high variance), which results in poor generalizability on the validation data [19], [32]. However, curating large scale training datasets of medical images with labels is challenging because of the tedious nature annotation processes and limited availability of domain expertise which makes the large scale data annotation expensive and time consuming, and fundamentally limits building effective medical imaging models across varying clinical use-cases.

Transfer learning using fine-tuning is one of the most popular strategies to address training with limited data in the radiology where the model is pretrained on a larger dataset (often with generic images, e.g. ImageNet) using supervised methods after which the model weights are fine-tuned for the target clinical domain with limited data [31]. The underlying assumption is that the supervised pertrained task and target task have significant similarity and thus, the low-level learnt features are common for both. Given the scarcity of labeled radiology images datasets, it is still common practice to pre-train medical imaging models using natural images of ImageNet dataset even though they do not share low-level visual features. However, recent studies show that pretraining on radiology images has huge potential [30]. Most medical imaging models under-perform on generalization which could be based on the fact that supervised pretraining paradigm encourages the model to mainly learn the highly correlated features with the specific labels from the larger dataset rather than general feature representation. Thus, most medical imaging models under-perform on generalization which could be based on the fact that supervised pretraining paradigm encourages the model to mainly learn the highly correlated features with the specific labels from the larger dataset rather than general feature representation.

In contrast to fine-tuning, Self-supervised Learning (SSL) is a process of training models to learn meaningful generic representation using unlabeled data and create foundation models [2] with the potential to transfer their knowledge of a variety of downstream target tasks - even when the pretraining task and target tasks are not similar [5], [7], [13], [16], [22], [23], [25], [26], [33]. Such models are used to learn image representations, encoding fundamental visual characteristics of the imaging data, and are tested for downstream prediction tasks like object detection on datasets like ImageNet. Simultaneously, complex vision-language models like CLIP [16], DALL-E [25], FLAVA [26], and Socratec models [34] were trained in a self-supervised way for understanding correlation in multi-modal data, and enabling reasoning across multiple modalities, and even applied in generation tasks. SSL is also being popular for radiology image analysis [3], [6], [20], [27]; however different from natural images, self-supervision using radiology images needs targeted pretraining using unlabeled medical images thus understanding the effects of various self-supervision strategies are important to plan experiments in advance and reduce unnecessary waste of computation resources and time of training. Although there exists a “gap” in literature since no study exists that benchmark SSL strategies for medical images.

For two popular medical image interpretation tasks (classification and segmentation), we designed encoder based self-supervised models and thoroughly benchmark a variety of self-supervision strategies for CT imaging data, i.e., i) masked area prediction, ii) next slice prediction, iii) rotation prediction, iv) flip prediction, and v) denoising. Our experiments covered the quality assessment of weights estimated by these strategies in terms of performance boost gained by downstream models when initialized with these weights. Additionally, performance disparities of AI models based on sensitive patient attributes is a major source of concern in wide-spread adoption of deep learning models [4], [14], [15], [28]. Image processing models for radiology images often learn patient attributes like race and use them as “shortcuts” for diagnosis and detection [1]. This behavior often leads to bias in models’ performance, especially against historically under-served populations. We also quantified knowledge of patient sensitive attribute seeping into self-supervised per-training as this bias may be also introduced into any downstream models built by initializing weights from self-supervised training.

As case-study, we focused on the commonly performed imaging modality of chest computed tomography exams. Potential downstream applications for chest CT are vast including segmentation of multiple organs like lungs and heart and/or segmentation of lung nodules, and detection of various abnormalities, such as pulmonary embolism, COVID-19, pneumonia, and lung cancer, coronary calcium scoring. Many of these tasks have been released with datasets as multiple public challenges. To counteract scarcity of annotated data at individual institutions, data in challenges are often pooled from multiple healthcare organizations and can make large, annotated sets available for relevant tasks like PE detection and lung nodule segmentation. However, this approach is not scalable for producing annotated sets for all relevant applications outside of the intended target (e.g., using publicly available PE dataset for cardiac segmentation).

On the chest CT use case, we evaluated the efficacy of self-supervised training on two separate clinical tasks (pulmonary embolism (PE) detection, lung nodule segmentation) and compared them to state-of-the-art models (SOTA) where we pre-train the CNN backbone using self-supervision with unlabeled private dataset and fine-tuned with limited task-specific public datasets. For baseline comparison, we also trained the same CNN backbone architecture without initializing with the pre-trained self-supervision weights.

## II. Materials and Methods

### A. Cohort Description

With the approval of the Institutional Review Board (IRB) for waiver of informed consent (IRB reference no. 21-005930), approximately 67,309 chest CT exams from more than 20k patients conducted between 2010 and 2022 were collected from various geographically disparate sites of our multi-site healthcare organization. Axial views of these studies were extracted and clipped to the soft-tissue window (50, 350) HU. Minimal filtering was done to preserve visual variance of the data to be generalizable for various tasks. Exams with slice thickness of more than 3mm were discarded. Table I shows salient characteristics of these exams.

**TABLE I:** Characteristics of all cohorts: self-supervised training and downstream target tasks: RSNA PE and Lung nodule segmentation (LIDC-IDRI).

|  | Self-supervised Training Data | RSNA PE Challenge | LIDC-IDRI |
| --- | --- | --- | --- |
| Dataset (volumes, slices) | 67309, 17509671<br>Slice thickness<br>≤3 mm:<br>51525, 15333281<br>Contrast-enhanced:<br>37709, 11324045 | 7279, 1790594 | 1012, 10005 |
| Findings | Lung related findings:<br>46% volumes<br>Cardiovascular findings:<br>12% volumes | PE:<br>2,211 volumes | Cancer:<br>388 volumes<br>Ambiguous:<br>547 volumes<br>No cancer:<br>394 volumes |
| Labels | – | PE:<br>96540 slices | Malignancy scores:<br>1: 923 slices<br>2: 1114 slices<br>3: 3473 slices<br>4: 2126 slices<br>5: 2369 slices |
| Patients | 19450 | – | 840 |
| Patient Age (mean +/- std) | 63.4+/-15.1 years | – | – |
| Manufacturer | SIEMENS: 31,089<br>GE: 9,696<br>TOSHIBA: 9,207<br>Cannon: 1,542 | – | – |

### B. Model Design

We experimented with two architectures for self-supervised learning (Fig 1); i) autoencoder architecture with encoder and decoder modules for image reconstruction based learning, and ii) encoder-only or classifier-type architecture for self-supervision through categorical labeling based learning. Encoder-decoder architecture was inspired by the vector quantized (VQ) autoencoder approach which maintains a code book of base vectors and weight vectors used to represent encoded input as a combination of base vectors [29]. Hence, the output of the encoder is discrete rather than continuous, and this approach is known for avoiding “posterior collapse” challenge where a powerful decoder ignores learned posterior. Our encoder model design included initial convolutional layers for image size compression and then a set of residual layers. Decoder was designed as a reverse of the encoder model to restore the original input image size. We designed our encoder-only or classifier-type architecture closely following the architecture of the encoder of our encoder-decoder type model. Compression and classification layers were appended to the encoder architecture to predict categorical self-supervision labels, e.g. rotation and flip prediction.

**Fig. 1:**
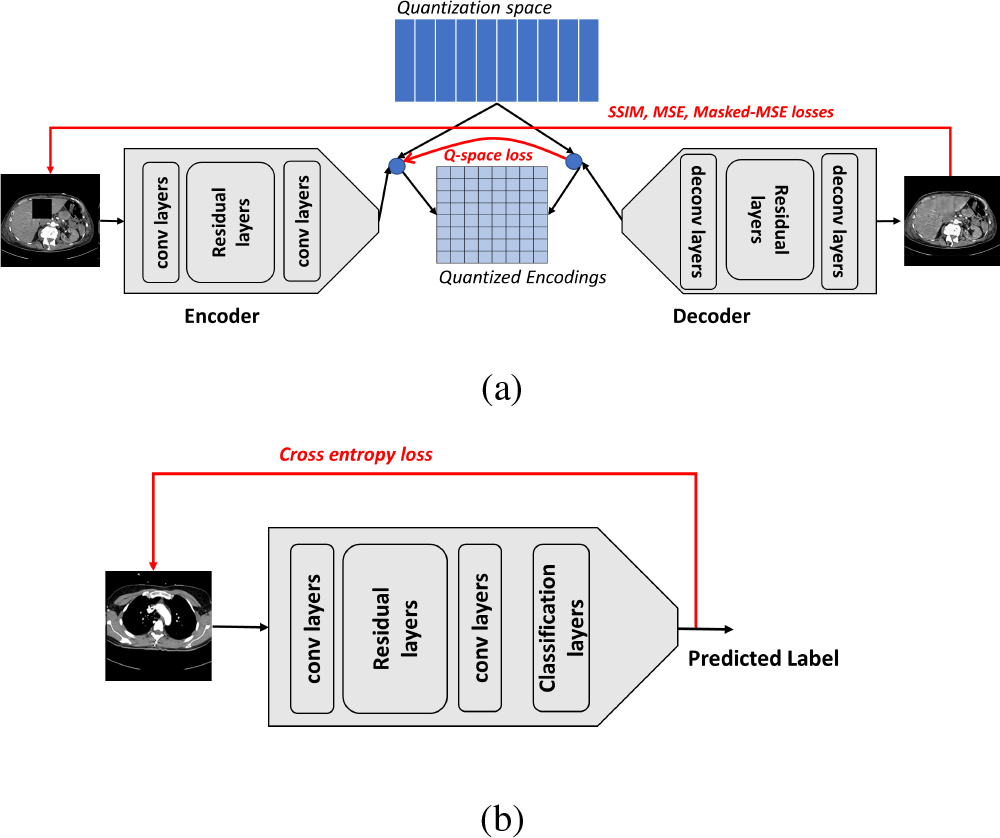
Self-supervised model architectures using VQVAE: (a) Encoder-decoder architecture, and (b) Encoder-only or classifier architecture.

Recent encoder-decoder architectures rely on letting the decoder obtain multi-scale cues from the encoder based on the skip connection; however, to ease the fine-tuning of the vector space for the target task [12], we purposefully avoided residual connections between encoder and decoder to force the model to return one compressed but informative vector in response to an input that is sufficient for producing decoder output (Figure 1a). The model consisted of approximately 100 million trainable parameters and was designed to encode and decode 512x512 pixel CT slices which is a common dimension of CT images. We designed a composite loss function with reconstruction loss (mean squared error (MSE)), reconstruction loss in quantized space, reconstruction loss of masked region, and negative of structural similarity index measure (SSIM). While MSE focuses on quantifying error in a reconstructed image (output image) compared to a reference image (groundtruth image), SSIM focuses on degradation of structural information in the reconstructed image when compared with the reference images. Classifier-type model is trained using straightforward cross-entropy loss as MSE and SSIM type losses are only applicable to image reconstruction model (Figure 1b). Self-supervised taining code is made available at https://github.com/amaratariq/FoundationChestCT.

### C. Strategies for Self-supervised Learning

We benchmark various self-supervision tasks for encoder backbone pretraining. Fig. 2 shows input-output pair selection for all approaches.

**Fig. 2:**
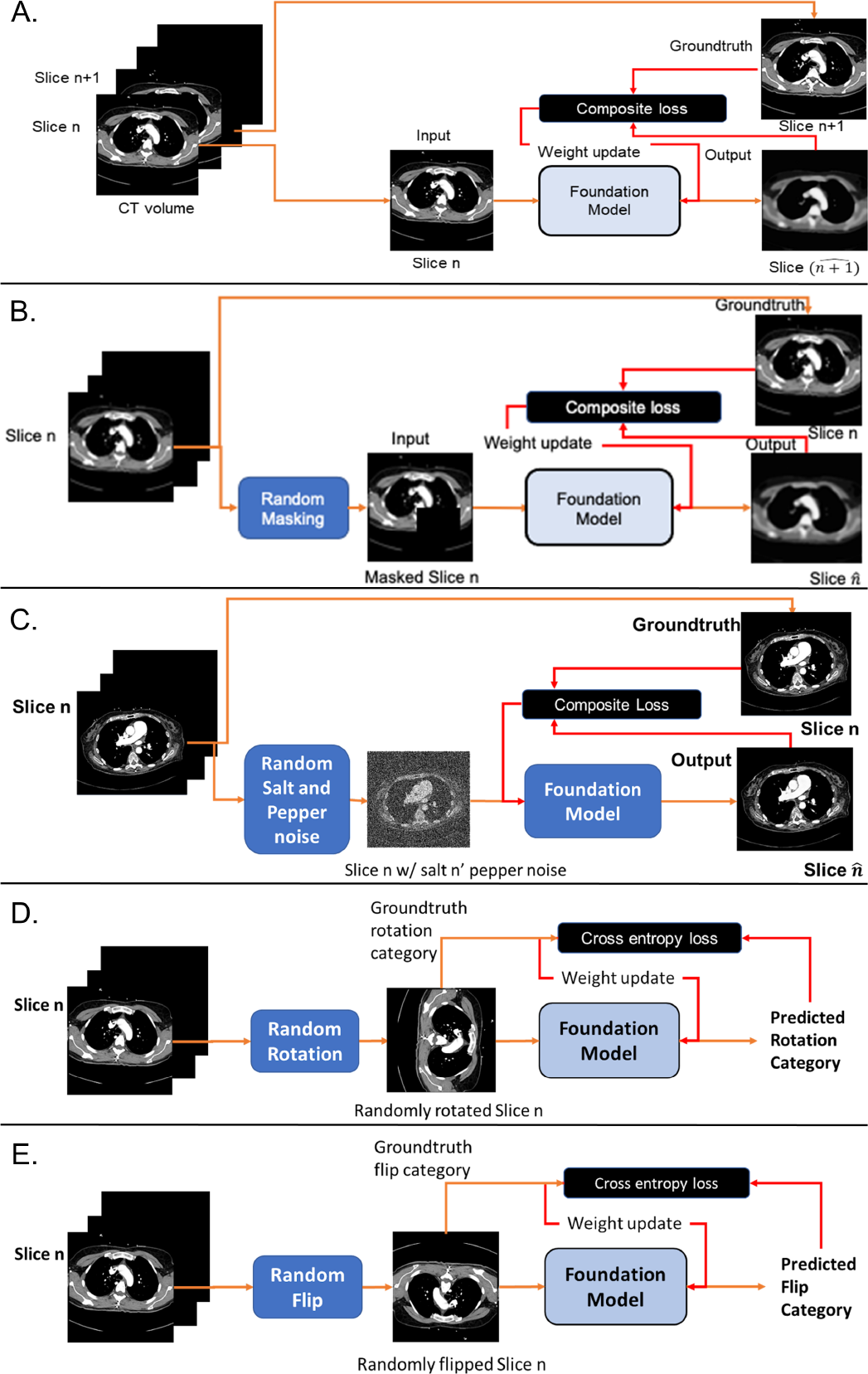
Self-supervision schemes for self-supervised model training – (A) next slice prediction; (B) masked region prediction; (C) Random salt and pepper noise removal (D) rotation prediction, (E) flip prediction

### 1) Next slice prediction

We leverage the 3D nature of CT volume and experimented with self-supervision through next slice prediction. Given the nth slice of the CT volume, the model was trained to generate the n+1th slice of the same CT volume under this self-supervision paradigm. Quality of generated output was measured by its similarity to the original *n* + 1^*th*^ slice of the volume. With *x* and *y* indicating input and output of the model, and *x*_*c*_ and 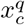 indicative compressed representation of *x* generated by *y* encoder and quantized representation of *x*_*c*_ generated by quantization layers, the following equation described the composite loss function for the model.

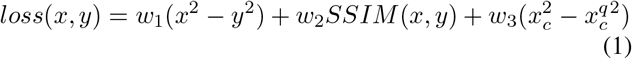

Weights *w*_1_, *w*_2_, and *w*_3_ can be finetuned as hyper-parameters.

### 2) Masked image region prediction

We also attempted self-supervision through randomly masked regions of the input slice and then training the model to re-generate the unmasked region. Performance of the model was judged by similarity of the output with the original unmasked input slice as well as model-generated patch for the masked input region and the original image patch for the same region. The loss function can be described by the following equation after denoting input masked region as *x*_*m*_ and model-generated patch for masked region as *y*_*m*_.

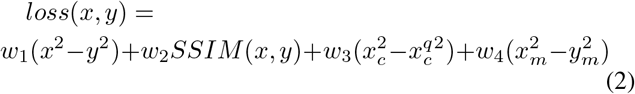

This self-supervision target was made progressively harder by masking larger and larger image regions, starting from masking of 5% of the total image area to masking of 20% of the total image area. Model trained by masking 5% image area was used to initialize the model trained with 10% image area masked, and so on. As the self-supervision target was made more complex, the model was initialized to take advantage of its earlier knowledge obtained under relatively easier self-supervision targets.

### 3) Denoising/Noise Removal

We experimented with self-supervised talk of noise removal, particularly for random salt and pepper noise at 50% level, i.e., noise was added to approximately half of the pixel with noisy pixels being randomly turned on (white or salt) and off (black or pepper). The model was trained to remove this noise and recover the original image. Composite training loss included mean squared error between predicted/denoised and groundtruth slice and SSIM, in addition to MSE in quantized space.

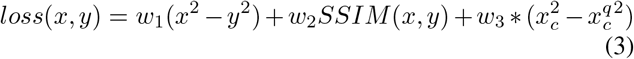

Predicted and groundtruth images are denoted by x and y respectively while xc and xcq represent compressed image and compressed image in quantized space, respectively. Weights w1, w2, and w3 can be finetuned as hyper-parameters.

### 4) Rotation prediction

We trained classifier-type self-supervised model architecture through prediction of categorical labels for CT slices where labels were set in self-supervised manner. First, we randomly rotated CT slices by 0, 90, 180, and 370 degrees. The self supervision learning task was set to predict the rotation category with the last classification layer designed for four-way classification. Cross-entropy loss used for model training can be described by the following equation such that *y*_*i*_ represents with *i*^*th*^ rotation category (set to 1 only if the input image *x* belongs to *i*^*th*^ category), and *p*(*y*_*i*_) represents the estimated probability of ith rotation category.

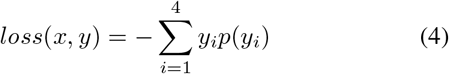

### 5) Flip prediction

We trained another classifier-type self-supervised model for flip prediction such that input slice can be flipped (i) horizontally, (ii) vertically or (iii) not flipped at all. Classification layer was set for 3-way classification. Cross-entropy loss was used for model training.

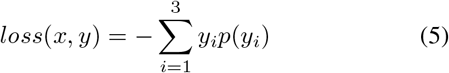

### D. Downstream Applications

#### 1) Pulmonary embolism detection

Radiology Society of North America (RSNA) conducted a competition in 2020^1^ to develop machine-learning algorithms to detect and characterize instances of pulmonary embolism (PE) on chest CT exams. The largest annotated PE dataset, consisting of approximately 12000 CT studies, contributed by five international research centers, and labeled by a group of more than 80 expert thoracic radiologists, was made publicly available for this competition (Table I). Given the importance and challenge related to the PE detection task, we employed this as one downstream application and compared PE detection models using pretraining weights from self-supervised models against the performance of the winning model of this challenge in comparative circumstances.

While availability of this multi-institutional dataset, approximately 1.8 million annotated CT slices, allows training of large, supervised models possible from scratch, collection of such large dataset is usually not feasible for every medical image processing model development. Therefore, we focused our evaluation experiments on a simulated but realistic scenario of availability of only a fraction of this large training data to analyze the effects of dataset size on model performance. We ran (a) PE detection model using pretraining weights, and the (b) RSNA challenge winning SEResNext model^2^ for different fractions of the data.

#### 2) Lung nodule segmentation

To assess the utility of self-supervised training for segmentation tasks, we chose lung nodule segmentation as our second task. The Lung Image Database Consortium image collection (LIDC-IDRI)^3^ consists of diagnostic and lung cancer screening thoracic CT scans with annotated lesions. Fifteen academic centers and imaging companies contributed to this set of 1018 cases (Table I). Annotations from four experienced thoracic radiologists were reconciled before rendering final annotations.

For this experiment, we used U-Net architecture given its vast application in medical image segmentation tasks [17]. We compared the performance of randomly initialized U-Net against U-Net with the “down” branch initialized by compression layers of our self-supervised model’s encoder architecture and weights. “Up” branch is initialized with the random weights similar to the baseline U-Net.

### E. Sensitive Patient Attribute Prediction

AI-based diagnostic models for radiology exams often exhibit strong predictive powers for patient attributes, and may find spurious correlation between these sensitive attributes and prediction tasks, resulting in patient attribute-based bias in their performance [1]. Since self-supervision is proposed as a building block for various downstream predictive models, we wanted to investigate the potential bias introduced by this foundational backbone. We extracted features for middle slices (as marked by the largest body cross section area covered) for a held out set of chest CT exams using self-supervised models trained with self-supervision techniques. By passing them through simple classification layers, we evaluate the relevance of these features to predict sensitive attributes related to patients -gender, race, and age group (bins of 10 years). As a comparative baseline, we build comparable predictors for raw images (middle slices) using an encoder model (from encoder-decoder architecture) appended with classification layers. We report predictive performance for patient attributes for features extracted by self-supervised models using all self-supervision techniques as well as raw slices.

## III. Results

### A. Comparative Performance of Self-supervision Tasks

We designed an experiment to test the performance of self-supervised models trained under all proposed self-supervision schemes, i.e., next slice prediction, masked region prediction (5%, 10%, 20% and 33% image regions masked), denoising, rotation prediction and flip prediction. Each model was tested on the same set of 128 CT volumes. Evaluation measures were based on self-supervision tasks. Image reconstruction based tasks (masked region and next slice prediction) were evaluated in terms of MSE (mean square error) and SSIM (structural similarity index matrix). For masked image region prediction, we also incorporated masked-MSE which is a measure of mean squared error between the original masked patch and the reconstructed patch. Label prediction tasks (flip and rotation prediction) were evaluated in terms of cross entropy loss (CE) and AUROC for prediction labels. Results of this experiment are presented in Table II.

**TABLE 2.**
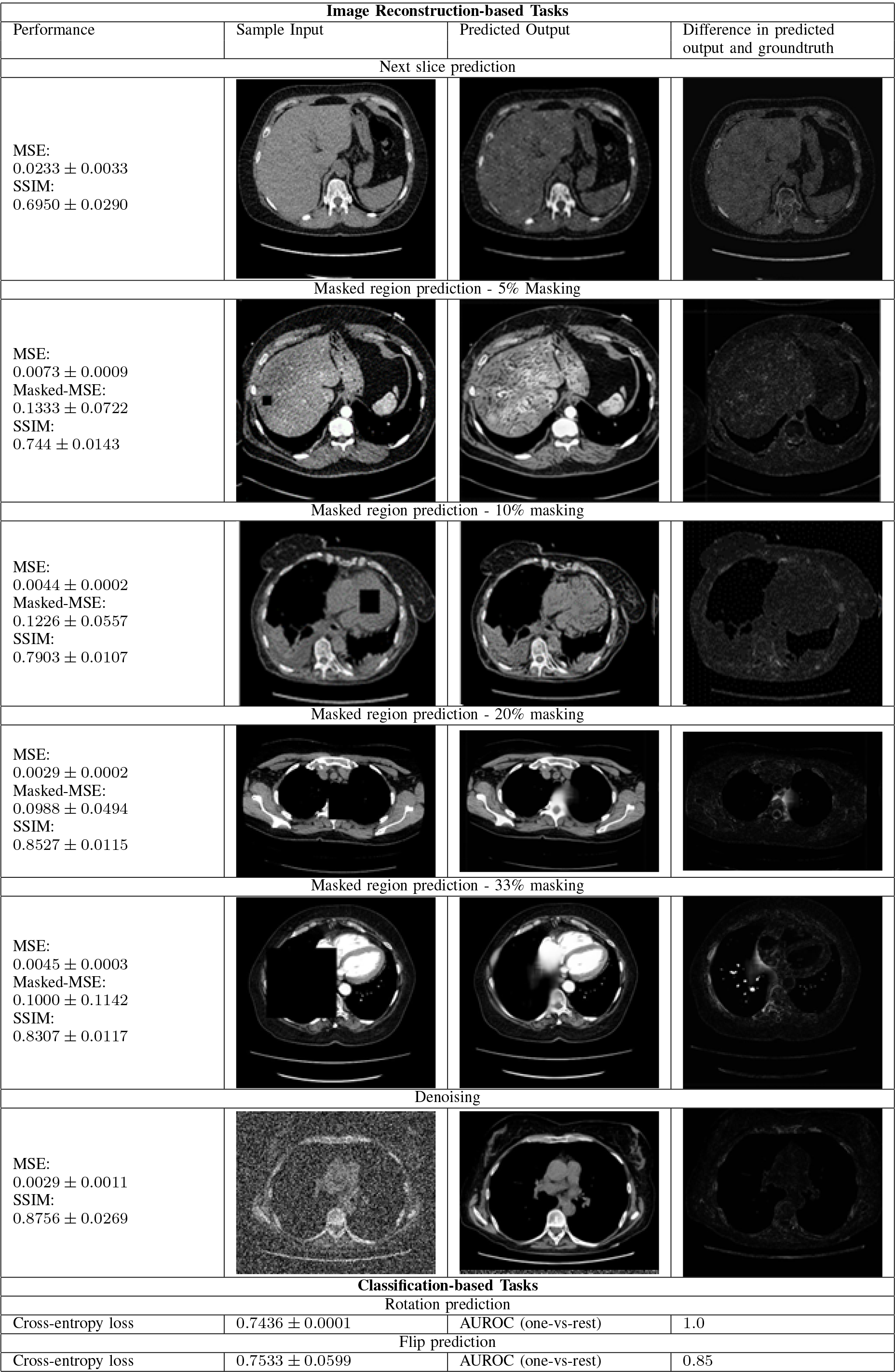
II: Performance of self-supervised training on 128 hold-out CT volumes.

| Image Reconstruction-based Tasks |  |  |  |
| --- | --- | --- | --- |
| Performance | Sample Input | Predicted Output | Difference in predicted output and groundtruth |
| Next slice prediction |  |  |  |
| MSE:<br>$0.0233 \pm 0.0033$<br>SSIM:<br>$0.6950 \pm 0.0290$ | | | |
| Masked region prediction - 5% Masking |  |  |  |
| MSE:<br>$0.0073 \pm 0.0009$<br>Masked-MSE:<br>$0.1333 \pm 0.0722$<br>SSIM:<br>$0.744 \pm 0.0143$ | | | |
| Masked region prediction - 10% masking |  |  |  |
| MSE:<br>$0.0044 \pm 0.0002$<br>Masked-MSE:<br>$0.1226 \pm 0.0557$<br>SSIM:<br>$0.7903 \pm 0.0107$ | | | |
| Masked region prediction - 20% masking |  |  |  |
| MSE:<br>$0.0029 \pm 0.0002$<br>Masked-MSE:<br>$0.0988 \pm 0.0494$<br>SSIM:<br>$0.8527 \pm 0.0115$ | | | |
| Masked region prediction - 33% masking |  |  |  |
| MSE:<br>$0.0045 \pm 0.0003$<br>Masked-MSE:<br>$0.1000 \pm 0.1142$<br>SSIM:<br>$0.8307 \pm 0.0117$ | | | |
| Denoising |  |  |  |
| MSE:<br>$0.0029 \pm 0.0011$<br>SSIM:<br>$0.8756 \pm 0.0269$ | | | |
| Classification-based Tasks |  |  |  |
| Rotation prediction |  |  |  |
| Cross-entropy loss | $0.7436 \pm 0.0001$ | AUROC (one-vs-rest) | 1.0 |
| Flip prediction |  |  |  |
| Cross-entropy loss | $0.7533 \pm 0.0599$ | AUROC (one-vs-rest) | 0.85 |

Results indicate that overall MSE and SSIM values improved when progressively increasing masked region size for masked image region prediction task up to 20% of the image regions. This trend can be attributed to the fact that each model in this progressive chain is initialized with the weights of the previous model (10% masked region prediction model with the weights of 5% masked region prediction, 20% masked region prediction model from 10% masked region prediction model). This allowed each model to retain knowledge learned in the past iteration of training and then learn further. However, improvement was not observed when masked image region size was increased from 20 to 33%. It may represent saturation of model’s learning capacity under the current selection of model and training dataset sizes. Better reduction is observed in overall MSE (39% improvement going from 5% to 10% masked region prediction, 35% improvement going from 10% to 20% masked region prediction) compared to reduction in masked region MSE loss (7% improvement going from 5% to 10% masked region prediction, 16% improvement going from 10% to 20% masked region prediction). Almost consistent improvement (7%) is observed at each step in SSIM values. For 20% masked region prediction, it is relatively straightforward for the model to reconstruct homogeneous image regions like patches masked from the liver. However, the model even learns to recreate complex shapes like vertebrae when masked (sample included in Table II) with relatively poor resolution at edges of the masked region. For 33% masked region prediction, the model seems to struggle with reconstructing when multiple organs are masked, reducing the context information available for reconstruction. Denoising also seems to be an effective self-supervision strategy. The model trained for denoising achieves as low MSE as the model trained with 20% mashed region prediction with even better SSIM (masked-MSE is not applicable to denoising model). For the next slice prediction model, the model has a relatively higher overall MSE highlighting the challenge of prediction of the next slice from a given slice, especially when slice thickness can be as large as 3mm. Label prediction tasks (flip and rotation prediction) performed comparably in terms of CE loss with rotation prediction achieving near perfect AUROC while flip prediction achieved high AUROC of 85%. Inherent horizontal symmetry in the axial view of chest CT made it difficult for the model to differentiate between horizontally flipped and non-flipped images, resulting in lower AUROC.

### B. Performance of Downstream Classification Task

Fig. 3 shows the results for PE detection. Performance comparison in terms of AUROC is presented for predictors initialized with self-supervised pre-training weights (masked regions prediction, next slice prediction, rotation and flip prediction) and state-of-the-art (SOTA) model. The 20% masked region prediction model was used as a representative of all masked image region prediction models as it outperformed 5%, 10% and 33% masked image region prediction models in self-supervised learning task evaluation experiments described in the previous section (Table 2). In general, predictors initialized with weights of the masked region prediction model outperform all other types of self-supervised pre-training weights across the board. They even outperform the SOTA model when only smaller training datasets were made available for training. Predictor initialized with masked regions prediction self-supervision weights outperformed SOTA for training data subsets as large as 770K slices, a dataset size which arguably too large for practical manual annotations. Model initialized with the weight of denoising based pre training also fared well achieving almost as high AUROC as model initialized with masked region prediction weights on dataset sizes as large as 115K. This result aligns with the results of self-supervision task based evaluation experiment where denoising based pretrained model achieved as low MSE as 20% masked region prediction based pretrained model. However, denoising based pre training weights did not perform as well as 20% masked regions prediction based pre training weights when even larger training datasets were used for PE detection model training. Both denoising and masked region prediction force the model to learn localized visual attributes through removal of noise from individual pixels and reconstruction of smaller image regions, respectively. This type of learning seems particularly useful for detection of small localized pathology of PE.

**Fig. 3:**
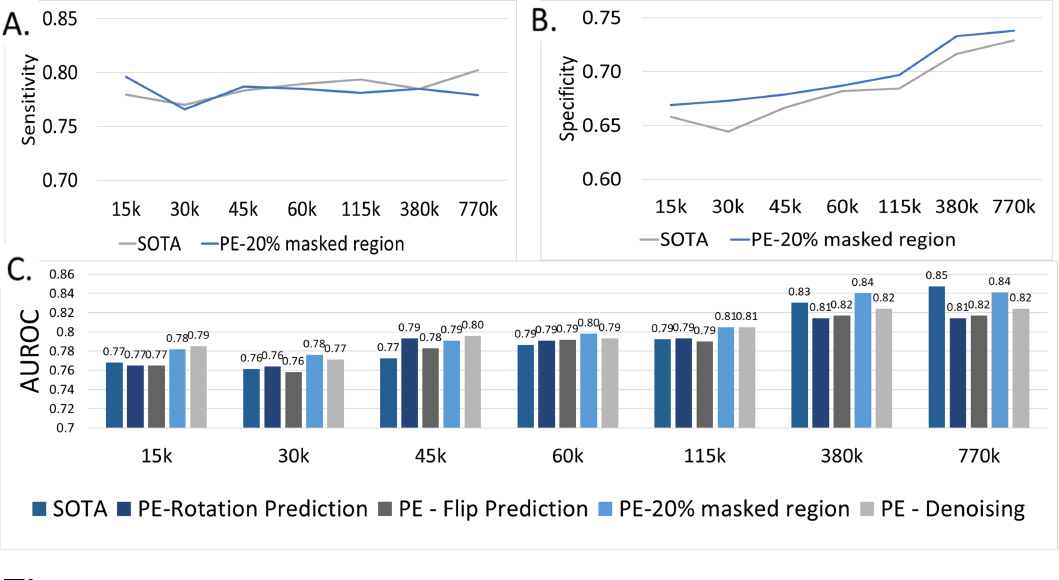
PE detection performance comparison between state-of-the-art PE detector (SOTA) and detectors using weights from self-supervised training - (A) Sensitivity and (B) Specificity reported for SOTA and the best performing predictor using pre-training weights (20% masked image region prediction), (C) AUROC reported for the SOTA and predictors with weights from all self-supervised tasks.

Predictors based on flip and rotation prediction training weights do show some gain compared to the SOTA model for training subsets of size up to 60K. However, their gains are much smaller compared to predictors using masked region prediction training weights. Again, this trend may be attributed to the fact that flip and rotation prediction based training does not force the model to learn localized visual features, instead focusing on over image orientation learning. We also plotted sensitivity and specificity values across a wide range of training data subsets for the SOTA and predictor using weights of 5% masked region prediction pre-training weights. While looking into sensitivity and specificity of these models, computed at the optimal probability threshold estimated by ROC curves, we observed that the detector using pretraining weights achieved consistently better specificity while keeping sensitivity value comparable to SOTA model - particularly when smaller subsets of training data (up to 770k slices) were used. Note that the supervised SOTA model was only tasked with learning one pathology (i.e., PE). On the other hand, the self-supervised models learned fundamental visual characteristics of CT data and then were able to transfer this knowledge, with only the architectural change of adding a fully-connected classification layer, for detection of a pathology with very small visual footprint even better than the supervised model narrowly focused on that one pathology.

We also compared the effectiveness of pretraining weights obtained by masked region prediction with varying mask sizes (5%, 10%, 20% and 33% of image size). Increasing the masked region size, especially beyond 20% of image size, does not seem to achieve consistent performance improvement in the downstream task of PE detection for smaller training dataset sizes of upto 60K (Figure 4). Some performance improvement is observed for even larger training dataset sizes. Performance of these pre-training weights on self-supervision tasks may offer a clue for explaining this trend. Under the current selection of dataset and model sizes, self-supervised training only seems to improve the pretrained model for up to 20% masked region prediction.

**Fig. 4:**
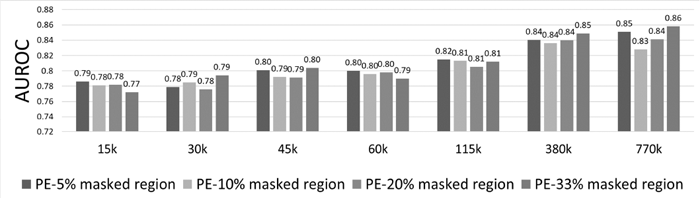
AUROC comparison between PE detection models built on top of encoders trained with self-supervised learning tasks of 5%, 10%, 20% and 33% masked region reconstruction

### C. Performance for Downstream Segmentation Task

Lung nodule segmentation is another challenging task requiring exact segmentation of a relatively small pathology of a lung nodule. U-Net is arguably the most robust medical segmentation model requiring relatively smaller training datasets. We focused our experiments on this architecture and compared two types of U-Net models; 1) U-Net initialized randomly (UNet-Random), and 2) U-Net with “down” formed of compression layers of self-supervised model encoder for all self-supervision tasks (U-Net-5% masked region, U-Net-10% masked region, U-Net-20% masked region, U-Net-33% masked region, U-Net-Denoising, U-Net-Next Slice, U-Net-Rotation, U-Net-Flip). Our hypothesis was that U-Net initialized with self-supervised pretraining weights has acquired some basic understanding of visual characteristics of CT scans through self-supervised training, and therefore should train faster for the task of segmentation for achieving comparable performance. The results supported our hypothesis for smaller training set sizes (horizontal axes of plots in Figure 5) consisting of up to 50% of all available training data containing about 100 studies with 3000 annotated slices with lung nodules. Again, 20% masked region prediction pretraining task was used as a representative of all masked region prediction models as it outperformed all other masked region prediction models in self-supervised task evaluation experiment (Table II). As the size of available training data was increased and training was run for larger and larger number of iterations, U-Net-Random tended to close the performance gap with U-Net initialized with pretraining weights, However, the experiment clearly established the benefits of using self-supervised pretraining weights under limited resource scenario. i.e., smaller amounts of annotated training data and limited computational time.

**Fig. 5:**
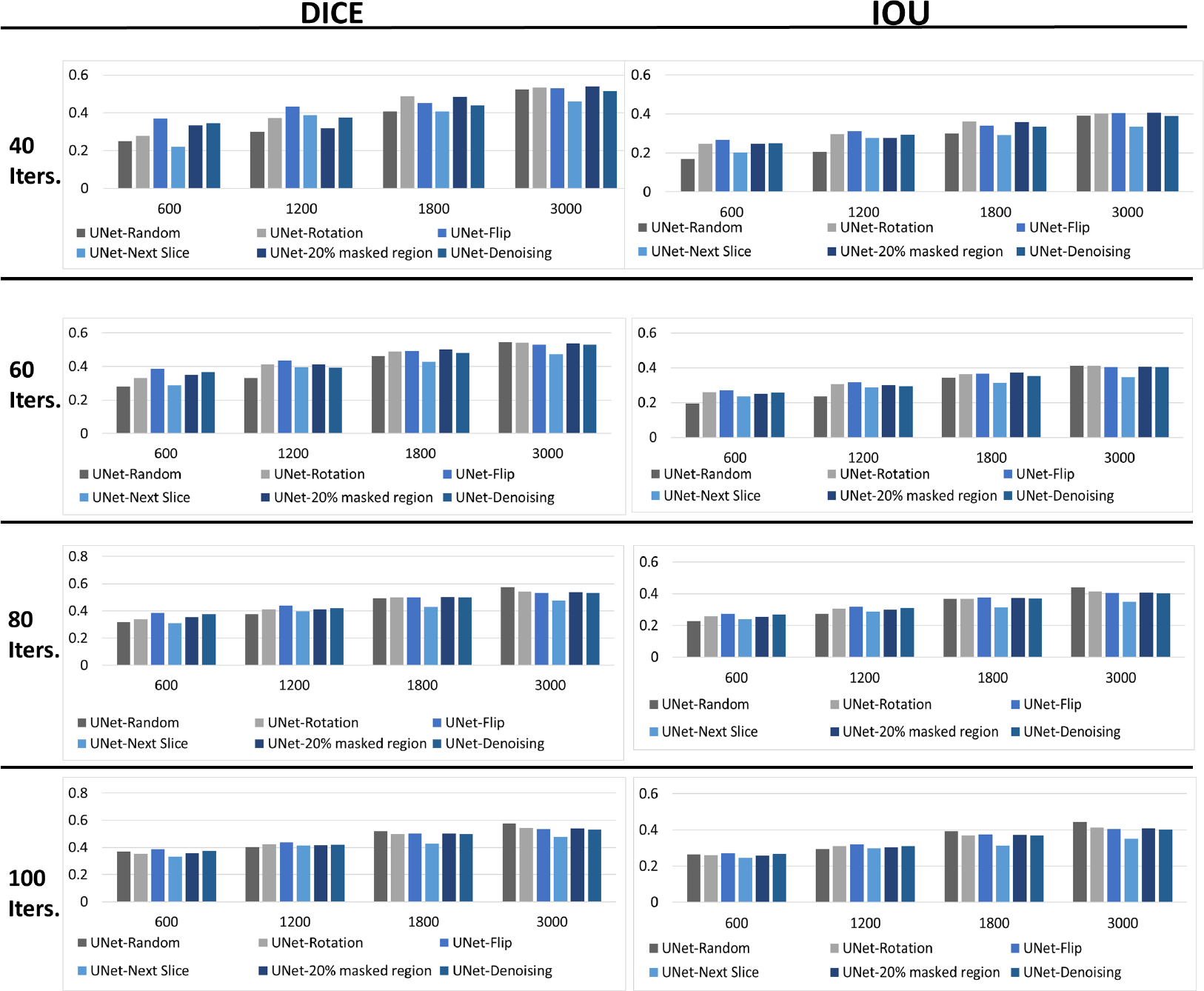
DICE and IOU comparison between UNet and UNet initialized by all self-supervised training weights for lung nodule segmentation over a wide range of available training dataset sizes and training iterations.

While masked region prediction seems to be a useful pretraining task for two very different downstream tasks, the nature of downstream tasks does affect the utility of pretraining weights. Tasks with extremely small visual footprint gain more from fine-grained pretraining tasks like masked region prediction. On the other hand, even global features learned by flip and rotation prediction are helpful for UNet to understand global visual structure (“down” branch of UNet) and use this knowledge for localized segmentation (“up” branch of UNet). Denoising based pretraining is also effective for both downstream tasks of PE detection and segmentation.

We also observed interesting patterns when we compared UNet initialized with pretraining weights of masked region prediction tasks with varying mask sizes (5%, 10%, 20%, and 33% image regions). Increasing masked region size generally improved the performance for downstream task of lung nodule segmentation when pretraining weights were used to initialize the “down” branch of UNet (Figure S1). However, this performance trend is stronger for larger training dataset sizes and numbers of training epochs. For very small training dataset sizes and numbers of epochs, no consistent gain is observed beyond 20% masked region prediction based pretraining. This result is consistent with performance of these pretraining weights for the downstream task of PE detection (Figure 4). This trend is also inline with performance of pretrained models on self-supervision tasks (Table II) where the model pretrained with 33% masked image region reconstruction failed to outperform 20% masked image region prediction model.

### D. Performance for Patient Attribute Prediction

While gender may have visible attributes in the chest CT, race and age represent a hidden sensitive attribute which is not straightforward to predict from the image. Well-known studies have shown that deep learning based image processing models for radiology images such as chest X-rays are astoundingly accurate at predicting race of the patient [14], [15], often resulting in damaging bias in performance of those models for minority groups. Predictive performance of image features extracted from all types of pretraining for all patients attributes are shown in Figure 6. In addition, performance of equivalent classifiers with no pretraining are also presented. For both gender and age prediction, performance gain of even the best performing self-supervision based features over no pretraining classifier is negligible. It appears that gender and age prediction gains little to no performance boost by using pretraining weights. It indicates that there is a minimal chance of self-supervision strategy introducing gender or age based bias in the downstream model performance beyond the bias acquired by the downstream model with no pretraining. Race prediction presents a somewhat interesting scenario. There is on average 10% performance boost gained when image features from self-supervised models are used, compared to the classifier with no pre-training. While gender and age have certain visual attributes in CT images and are thus easier targets to predict for image processing model, prediction of these target does not seem to need any help from pretraining based image features. On the other hand, race is a difficult prediction target with no well-defined visual attributes. Prediction of such a target is aided by the image features obtained through self-supervised training. This may increase the chance of race-based bias in the downstream models initialized with self-supervised pretraining weights, while gaining advantage in terms of down-stream task performance (better results with smaller amounts of training data and computational resources). However, race prediction performance for image features from all types of self-supervised image features is pretty even (AUROC between 0.72 and 0.74), except for the next slice prediction based model (AUROC of 0.68). This self-supervision strategy also shows the lowest performance gains for downstream tasks of PE detection and lung nodule segmentation, indicating overall poor quality of features obtained through such self-supervision. Overall, the chance of introduction of bias in downstream tasks seems to be equally strong for all otherwise effective self-supervision strategies.

**Fig. 6:**
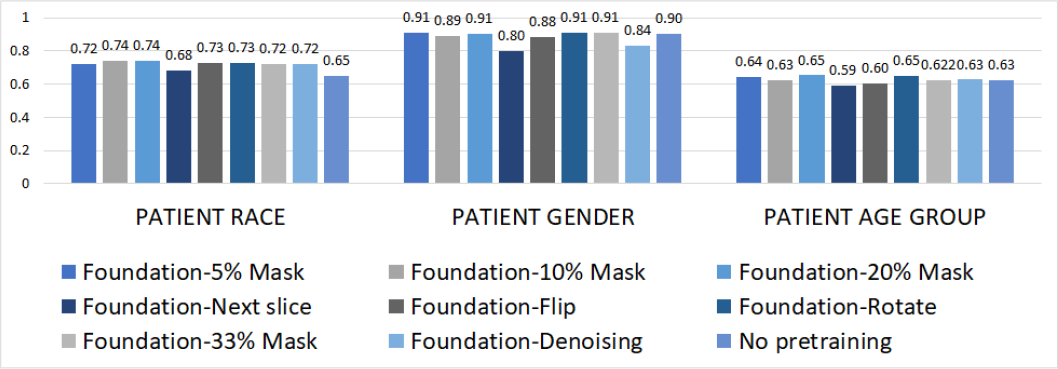
AUROC based performance comparison for patient attribute prediction

## IV. Discussion

To the best of our knowledge, we have presented the first comprehensive benchmark study of self-supervised learning strategies for chest CT exams for potential application in a variety of downstream tasks such as PE detection and lung nodule segmentation, in addition to evaluating the effects of self-supervision of patient attribute based bias.

Self-supervised learning strategies discussed in this work present a systematic solution to this challenge of generating task-specific large datasets by learning fundamental visual characteristics of CT from un-annotated CT volumes in self-supervised fashion and then transferring this knowledge to downstream tasks. Our experiments show that the downstream prediction models initialized with pretraining weights learn faster (saturates with fewer epoch) and outperform state-of-the-art supervised downstream classification and segmentation tasks when only small amounts of annotated training data is available. While nature of the downstream task affects the efficacy of pretraining weights coming from different types of self-supervised training, masked region prediction seems to be an overall effective method for pretraining for chest CT performing well for widely different tasks of PE detection and lung nodule segmentation. It is the best strategy for detecting small visual footprint pathology (e.g., pulmonary embolism).

In a practical scenario, users can directly download pre-trained weights and transfer them to other chest CT tasks, such as coronary calcium scoring. We have presented frameworks for using these weights for two vastly different models (CNN based classifier, and UNet based segmentation model). The significance lies in reducing the dependency in not only reducing the amount of annotations required but also reducing the dependency on having large dataset sizes, both of which may not always be practically possible.

In addition to the results we observed, we also noticed interesting behaviors during self-supervised training. In the self-supervised learning paradigms, which was built around next-slice prediction, the model primarily focused on compression rather than learning anatomy visible in slices. When tested on downstream tasks (Figure 3 for PE detection and Figure 5 for lung nodule segmentation), the performance of the downstream model using weights of this self-supervised training strategy was poor in comparison to models using weights of masked regions prediction pretraining. The performance gap was wider for smaller training dataset sizes. Categorical label prediction based self-supervision (rotation or flip prediction) seems to acquire knowledge of global visual characteristics rather than fine grained localized visual features, as indicated by the relatively poor performance of pretraining weights obtained by such self supervision on PE detection, but substantial performance gain obtained by the same weights when used to initialize “down” branch of UNet for lung nodule segmentation.

While PE detection and lung nodule segmentation experiments clearly establish the utility of pretraining weights obtained by self-supervision, their effect on potential introduction of bias in downstream models warrant separate investigation. We studied this effect by training patient attribute predictors on a small held-out set of patients with and without pretraining weights. Our experiments show that attributes with visible features like gender or age are easily learned by the predictors even without the help of pretraining weights, indicating minimal chance of introduction of additional bias through pretraining weights for these attributes. Prediction of attributes with no definitive visual features like race is improved with the use of pretraining weights. however, pretraining weights from all effective self supervision strategies seem to learn race features almost equally well. It is possible that bias based on hidden patient attributes may be present in pretraining weights when they are used to initialize downstream models. However, significant advantages in terms of performance boost for limited training datasets and computational resources are observed for downstream models when initialized with pretraining weights. We argue that such performance gain warrants the use of pretraining weights while highlighting the need for debiasing of downstream models. Vast amount of literature in the field of medical image processing is now focused on experimenting with training of image processing models with debiasing techniques [8]–[10]. Such techniques will be applicable even when the models are initialized with pretraining weights.

Given the potential, we hypothesized that self-supervised learning can allow a paradigm shift for AI model development, where many models across domains will directly build upon or heavily integrate self-supervised models. Self-supervised learning incentivizes homogenization where the same backbone architecture is repeatedly reused as the basis for diverse applications. Such consolidation allows centralization to concentrate and amortize community efforts (e.g., to improve robustness) on a specific model that can be repeatedly applied across applications to reap these benefits.

### Limitation

Within the scope of this study, we only experimented with a simplistic encoder architecture that suits a wide range of tasks – however we demonstrate the application for both the segmentation and classification tasks. As the first step, we studied self-supervised learning for 2D slices of CT volumes. In future, we plan to extend these experiments for 3D CT volumes. Our experiments concluded that 20% masking is the optimal self-supervision task for downstream targeted classification and segmentation. However, this experiment is restricted by the size of the pre-training data (67,309 volumes; 17,509,671 slices). More pretraining data may also help the model to achieve better reconstruction for the larger masked regions.

## Data Availability

All data produced in the present study are available upon reasonable request to the authors

## Footnotes

1 https://www.rsna.org/education/ai-resources-and-training/ai-image-challenge/rsna-pe-detection-challenge-2020

2 https://www.kaggle.com/c/rsna-str-pulmonary-embolism-detection/discussion/194145

3 https://wiki.cancerimagingarchive.net/pages/viewpage.action?pageId=1966254

## Notes

### Competing Interest Statement

The authors have declared no competing interest.

### Funding Statement

This study did not receive any funding

### Author Declarations

Ethical approval granted by Internal Review Board (IRB) of Mayo Clinic.

