## Supplementary Figure for "Self-supervised Learning for Chest CT - Training Strategies and Effect on Downstream Applications"

### I. Supplementary Material

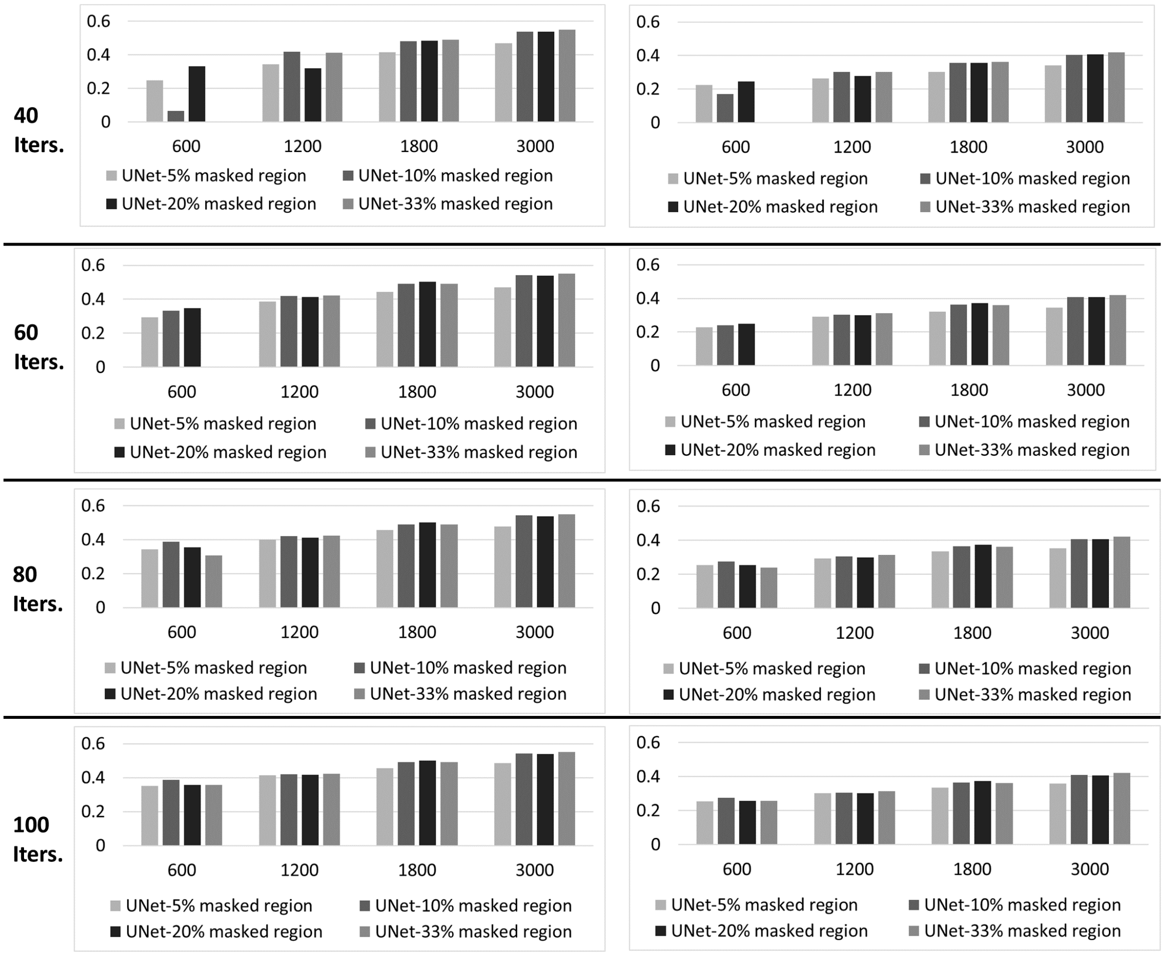


Fig. S1: DICE and IOU comparison between UNet using weights from self-supervised learning using 5%, 10%, 20% and 33% image masking
